# Perspectives from EPI personnel on immunisation programme performance in Kenya: a thematic, complex adaptive systems and leverage-points analysis

**DOI:** 10.64898/2026.09.26.26364071

**Authors:** Christine Karanja-Chege, Dan Odallo, Ambrose Agweyu, Rose Jalang’o, Fred Were, Michaël Boele van Hensbroek, William Ogallo

## Abstract

**Background:** Immunisation system performance depends on effective service delivery and the generation of high-quality data to support planning and decision-making. With the recent scaling down of external health funding, Kenya must identify sustainable strategies to strengthen the resilience of its immunisation programme.

**Methods:** We conducted a three-part qualitative study comprising thematic analysis, application of complex adaptive systems (CAS) theory, and the Meadows leverage-points framework. Thematic key personnel interviews were conducted with selected Essential Programme on Immunisation (EPI) personnel from 14 counties and sub-counties in Kenya to explore factors influencing immunisation system performance.

**Results:** Vaccine distribution, health workforce shortages and immunisation financing emerged as the dominant themes affecting immunisation performance. Analysis through a CAS lens revealed that reinforcing feedback loops were more dominant than balancing loops, suggesting that system dysfunctions were more influential than adaptive self-correcting mechanisms, resulting in a tendency toward negative system behaviour. Application of the Meadow’s framework showed that existing interventions were largely concentrated at lower leverage levels, which are more likely to generate short-term operational improvements, while higher-leverage interventions with the potential for long-term system transformation remained underutilised.

**Conclusion:** Systems-thinking approaches offer a powerful framework for understanding and improving immunisation system performance. They facilitate the identification of vulnerabilities and bottlenecks, while informing the design of higher-leverage interventions capable of producing sustainable and transformative improvements across the system.

## Background

Sustaining high and equitable immunisation coverage remains a priority under both the Kenya National Immunisation Policy [1] and the global Immunisation Agenda 2030 (2). In Kenya, coverage has risen steadily over the years, plateauing at around the mid-eighties mark, mirroring global estimates [3]. Despite this progress, significant sub-national disparities persist [4, 5], underscoring the need to better understand the underlying system-level and contextual factors influencing immunisation performance.

Effective immunisation service provision relies not only on the efficiency of service delivery but also on the availability of accurate data - both for estimating target populations and for assessing programme performance [6, 7]. Inaccurate denominators, vaccine dose recording errors and inconsistent reporting at subnational levels distort coverage estimates and obscure true service delivery gaps [8]. Such issues have been documented in several low-and middle-income countries, including Kenya, where health information systems continue to evolve [7].

Our team previously conducted a quantitative analysis examining key aspects of Kenya’s immunisation information and service delivery system. In that study, we explored the relationship between dropout rates, immunisation coverage and health facility density across all 47 counties (sub-national administrative units in Kenya). A sub-analysis of DTP1 and DTP3 vaccination trends from 2020 -2024 showed diverse patterns across counties. While the analysis revealed considerable inconsistencies in data metrics and inequities in service delivery, it could not fully explain the operational and contextual drivers of these patterns [9]. We therefore selected seven counties that exhibited particularly distinct trends for qualitative exploration.

In the present study we adopted a triangulated approach by eliciting insights from key personnel directly involved with immunisation programme implementation, thereby complementing the quantitative findings with qualitative evidence. Through thematic key personnel interviews with county and sub-county EPI staff, we explored how data-related and service delivery challenges are encountered and responded to in practice, and how the dynamic relationships among key operational factors influencing immunisation service delivery shape overall system performance. In addition, we examined leverage points within the system that can be used to design optimal interventions to address the existing systemic constraints.

We hope that findings from this qualitative study will provide critical insights to guide data-driven decision making and inform strategies for strengthening immunisation health system performance, thereby contributing to more equitable immunisation service delivery across the country.

## Methods

### Research design

The study employed a qualitative design with three analytic frameworks: Thematic, complex adaptive systems and leverage-points analysis.

Thematic analysis was done using a deductive coding approach and inductive identification of emergent sub-themes. These themes and sub-themes were translated into system constructs and the complex adaptive systems (CAS) framework applied to link the constructs into feedback relationships and system dynamics within the immunisation system. The CAS theory provides a framework for understanding systems composed of multiple elements that interact with each other within a dynamic environment and produce outcomes that cannot be explained by the individual components in isolation [10].

The Meadows’ leverage-points framework was subsequently applied to identify potential intervention points in the system to improve performance. Meadows proposed a hierarchy of twelve leverage points representing opportunities where interventions can be applied to alter the behaviour of complex systems [11].

### Tool development and data collection

The thematic key personnel interview guide (supplementary material 1) and deductive coding framework were informed by the WHO AFRO EPI operational guidance [12] and adapted using the WHO Health System Building Blocks framework [13].

The following seven factors were explored using the interview guide: vaccines and commodities procurement process, the distribution process, storage and cold chain equipment, health workforce, data management, communication, service delivery, and immunization financing at county and sub-county levels.

The interview guide was pilot-tested for flow and coherence with four nurses providing immunisation services in Nairobi County. They were not included in the study. Feedback from this process was used to refine the guide by reducing ambiguity, improving the clarity of the questions and adjusting the estimated interview duration.

### Participant recruitment and selection criteria

We purposively selected 14 respondents from 7 selected counties that demonstrated distinct patterns in the preceding quantitative analysis including immunization coverage, dropout rates and health facility density [9]. The selection criteria are outlined in table 1.

Data saturation was assessed iteratively during data collection. By the twelfth interview, there was no emergence of new themes, and the final two interviews served to confirm and enrich existing themes. We therefore determined that theoretical sufficiency had been attained for the purposes of analysis.

**Table 1:** County selection Criteria based on Immunisation coverage, dropout rates and health facility density.

| **County** | **Immunisation coverage** | **Trend** | **Dropout rates** | **Health facility density** |
| --- | --- | --- | --- | --- |
| County A | High coverage | stable trend | high dropout rates | high |
| County B | Low coverage | stable trend | low dropout rates | low |
| County C | High coverage | stable trend | negative dropout rates | high |
| County D | Low coverage | downward trend | low dropout rates | high |
| County E | Cyclic coverage pattern | stable trend | low dropout rates | low |
| County F | low coverage; | upward trend | moderate dropout rates | low |
| County G | Low coverage | stable trend | high dropout rates | high |

Source: Karanja-Chege et al, 2026 [9]

**Source:** Karanja-Chege et al, 2026 [9]

**National mean coverage (85%):** high coverage; ≥85%; low coverage: <85%

**WHO Dropout rate thresholds:** high > 10%; moderate: 5-10%; low: <5%; negative dropout rates: negative values

**National average Health facility density (2.89/10000 population):** low: <2.89; high ≥2.89

From each county, we interviewed the county EPI manager and a sub-county EPI focal person or nurse. They read and each signed the informed consent form (Supplementary material 2). The de-identified and anonymised participant details are shown in table 2

**Table 2:** Participant Characteristics (n=14)

| Respondent Number | County/Subcounty | Role |
| --- | --- | --- |
| P1 | County A | County EPI coordinator |
| P2 | County B | County EPI coordinator |
| P3 | County C | County EPI coordinator |
| P4 | County D | County EPI coordinator |
| P5 | County E | County EPI coordinator |
| P6 | County F | County EPI coordinator |
| P7 | County G | County EPI coordinator |
| P8 | Sub-county A | Sub-county EPI coordinator |
| P9 | Sub-county B | Sub-county EPI coordinator |
| P10 | Sub-county C | Sub-county EPI coordinator |
| P11 | Sub-county D | Sub-county EPI coordinator |
| P12 | Sub-county E | Sub-county EPI coordinator |
| P13 | Sub-county F | Sub-county EPI coordinator |
| P14 | Sub-county G | Sub-county EPI coordinator |

### Data collection

Once consent was obtained, we used a thematic key personnel interview guide (supplementary material 1) to conduct online interviews with each of the 14 EPI personnel. We conducted nine of the interviews via an online video platform. This was recorded by manual note-taking as well as an AI transcribing tool. All the meeting transcripts were collated with the interviewer’s notes and uploaded to a secure desktop folder accessible only to the investigators. We made direct phone calls to five interviewees where internet connectivity was unreliable, and took detailed field notes. The consistency of themes across both interview modalities suggests that data quality was not substantially compromised.

### Data Verification and analysis

The lead investigator (CK) and a co-investigator (DO) systematically reviewed the transcripts, to ensure accuracy and completeness. This process involved reflecting on the data collection process and refining the verbatim transcripts to maintain fidelity to the participants responses.

We conducted deductive thematic analysis of the seven pre-defined domains, following the approach proposed by Braun and Clarke [14]. The lead investigator (Ck) reviewed the de-identified interview transcripts, developed codes and identified emerging sub-themes. A co-investigator (DO) independently reviewed the coding and thematic structure. Discrepancies between the two reviewers were resolved through discussion until consensus was reached. We visualised the validated themes on a thematic map. Microsoft Copilot (Microsoft Corporation) was used as an analytical support tool during the coding process to assist with organisation of de-identified transcripts [15].

We translated the themes to system-level constructs and applied the complex adaptive systems theory to examine the dynamic interactions among them. This process enabled the identification of feedback loops illustrating how certain factors contributed to the deterioration of immunisation system performance while others supported self-organisation and system resilience. Each proposed feedback loop was validated against the original interview excerpts to ensure empirical grounding.

We further applied the Donella Meadows’ leverage-points framework to map existing interventions and identify potential high-impact intervention opportunities within the immunisation system. Meadows identified 12 hierarchical leverage points where application of an intervention within a complex system can produce a massive, transformative change [11].

We synthesised and reported these findings in a narrative write-up. All analyses were performed and visualised in Python version 3.13 (matplotlib and NetworkX) [15].

We reported our findings in accordance with the Consolidated Criteria for Reporting Qualitative Research (COREQ) checklist [16].

### Ethics Statement

Ethical approval was obtained from the Kenyatta University Ethical Review Committee (Reference number PKU/2575/11701). Written informed consent was obtained from all participants prior to data collection. Participant information was de-identified and anonymised to maintain confidentiality, and participation was voluntary.

### Reflexivity and Positionality Statement

The lead investigator (CK) is a Kenyan paediatrician and global child health researcher with experience in immunisation systems research. Her professional background facilitated familiarity with immunisation service delivery but may have influenced the interpretation of participant narratives. The interviewees were informed that their responses would be anonymised and throughout the analysis reflexive discussions were conducted within the research team to challenge assumptions.

## Results

### Thematic analysis

There were seven pre-set themes from which emerged 20 sub-themes as shown in the coding framework summary in supplemetary figure S1.

### Theme 1: Vaccine Procurement and distribution

All 14 transcripts confirmed a centralised national procurement model where vaccines are funded and procured by the government through the National Immunisation Programme. The supply chain pathway: National Depot -> Regional Store -> County Store -> Sub-County Store - > Health Facilities. We identified 3 emergent sub-themes as follows:

### Transport as a bottleneck in distribution

Transport emerged as the single most significant constraint affecting vaccines distribution across all 14 counties and sub counties. It manifested as: insufficient numbers of vehicles, lack of fuel, competing demands for vehicles and challenging terrain.

Respondent P1: “*Difficult terrain and poor roads especially after rains.*” Respondent P5: “*Only 3 vehicles; vehicle breakdown; low fuel allocation.*”

Respondent P9: “*Facilities use public transport and the conditioned ice-packs are not always adequate for far flung areas.*”

### Vaccine formulation and switch challenges

Several respondents noted that vaccine switches (e.g. rotavirus vaccine switch) and newer formulations create supply disruptions and vaccine vial monitor (VVM) stability concerns.

Respondent P8: “*Newer vaccine formulations like rota and IPV [inactivated polio vaccine] change their VVMs too quickly.*”

Respondent P2: “*Some vaccines have been out of stock e.g. rota due to the switch.*”

### Innovative distribution solutions

Drone delivery (Zipline): Used in County E for hard-to-reach areas and emergency re-stocking during outreaches – Respondent P12.

Last Mile Delivery project: National-level and partner supported initiative delivering vaccines directly to facility doorsteps – Respondent P2, Respondent P9.

Redistribution between facilities: Moving stock from low-volume to high volume facilities to manage shortages – Respondent P2, Respondent P4, Respondent P12.

### Theme 2: Cold Chain storage and equipment

All counties reported adequate and improved cold chain capacity. However, maintenance and spare-parts were the biggest challenges.

### Security threats to cold chain infrastructure

This was a unique finding specific to a pastoral county where solar cold chain equipment vulnerable to theft.

Respondent P3: “*theft of solar panels through break-ins.*”

### Maintenance and spare-parts challenges

There is no dedicated funding for equipment maintenance, transport of biomedical engineers and purchase of spare-parts.

Respondent P7: “*No allocated budget for maintenance*”

### Theme 3: Health Workforce

Across all the transcripts, severe workforce shortage was a consistent finding. It was described as a root cause and amplifier across the other themes.

### Service disruption when sole nurse is absent

When the only nurse running the facility is absent or away on training, the facility is closed entirely and patients are redirected to distant alternatives.

Respondent P9: “*When the nurse is away, patients should be re-directed to other facilities which may be too far.*”

### Private sector staff drain

Respondent P13: *“Attrition of EPI trained staff due to more lucrative terms in private facilities.*”

### Supervision and mentorship deficit

Supervision support is used to fill training gaps and is not adequate. It is constrained by lack of transport and funding.

Respondent P13: “*Supervision support is not adequate to offer mentorship to fill the gap.*”

Respondent P4: “*Support supervision is integrated with other programs due to funding challenges.*”

### Theme 4: Immunisation financing

Financing was a cross-cutting constraint underpinning nearly all other challenges. It also emerged that immunisation is not prioritized in county health budgets.

### Partner dependency for core operations

Operational activities that should be government funded rely heavily on partner support (WHO, UNICEF, World Bank, Aga Khan).

Respondent P12: “Aga khan recently supported HPV outreach activity.” Respondent P10: “There is a designated vehicle provided by WHO.”

Social Health Authority (SHA) as an emerging financing mechanism. Respondent P9: “SHA now provides funds to buy syringes from MEDS [Mission for Essential Drugs and Supplies].”

Respondent P12: “There is now SHA allocation for supervision activities.”

### Integration with other programs as a survival strategy

Respondent P12: “*To cope with funding constraints, outreach activities are integrated with better funded programmes.”*

### Theme 5: Data and records

Data is managed through a dual manual-electronic system. Facilities use paper-based tools for recording immunisation data, while counties and sub-counties use digital platforms for data aggregation and ordering.

### Staffing as a data quality determinant

Data quality is directly correlated to workforce capacity – single-nurse facilities produce poorer data because the same person providing care must also document and report.

Respondent P1: “*Staffing shortages at facilities cause reporting delays when that person is absent.”*

Respondent P13:”*HRIOs [Health Records and Information officers] are busy with non-EPI indicators — the better funded programmes like HIV/TB.”*

### Emerging digital health technologies

New digital platforms are being piloted alongside E-Chanjo.

Tibabu platform: Being rolled out in County C (Respondent P3); piloted in County E with data losses noted (Respondent P5).

TaifaCare: Piloted in County D for defaulter tracing (Respondent P4)

### Stock-outs of data collection tools

Respondent P3: *”Shortage of manual registers; internet challenges.”*

Respondent P11: “*Stock outs of manual data collection tools — county does not support transport to go collect.”*

### Theme 6: Communication

Communication in the immunization program follows a standardized cascade: National -> County Director of Health -> County EPI head -> Sub-County EPI coordinator -> Facilities -> Community. Digital channels (email, phone messaging) are the primary mode for formal communication.

### Vaccine hesitancy is a major communication challenge with distinct patterns

i) HPV vaccine linked to fertility risks

Respondent P14*: “Rumours that HPV vaccine is laced with FP.”*

ii) Religious reasons for refusal

Respondent P4: *“Catholic schools and FBOs [Faith Based Organisations] refuse HPV.”*

iii) Resistance in the middle class/urban population

Respondent P2: *“Why this time? Why our children?”*

iv) Community-specific hesitancy

Respondent P13: *“Somali community refuses MR [measles-rubella] but accepts TCV [typhoid conjugate vaccine].”*

v) Health worker hesitancy

Respondent P4: “Some HCWs [Health care workers] are hesitant about HPV[human papilloma virus] and malaria vaccines.”

### Ministry of Education (MoE) to Ministry of Health (MoH) coordination gap

The lack of a harmonised strategy between MoE and MoH specifically undermines school-based HPV vaccination.

Respondent P7: “M*oE and MoH do not have a harmonised strategy — a lot of back and forth leading to delays.”*

Respondent P9:*”HPV School programme — refusal by Catholic schools”*

Peer-led and Community-based mitigation strategies Respondent P11:”Somali peer educator talked to them.”

Respondent P3 *”Health talks in local radio stations eg Serian FM.”*

Respondent P1:*”Continuous dialogue, community education, and the community health strategy.”*

### Theme 7: Service delivery

Immunisation service delivery across the 14 counties and sub-counties operates through three modalities: routine health facility based (main), outreach (determined by availability of funding), school-based (for HPV vaccination).

### Cross-border service delivery challenges – unique to a border sub-county

Respondent P11:”*Sub-county B borders Uganda; cross over of clients from and to either side; misclassification and loss to follow up — defaulter tracing difficult.”*

Respondent P11: *Mitigation: “Vaccination point at port-health to cover these clients.”*

### Community Health Strategy as a performance driver

The Community Health Strategy: Community Health Assistants (CHAs) -> Community Health Promoters (CHPs) emerged as the strongest positive service delivery factor, enabling defaulter tracing, community mobilization and demand creation.

Respondent P1:*”Strong community health strategy and active community health volunteers/CHP system for defaulter tracing and community mobilization, which likely contributes to County A’s good performance and low dropout rates.”*

Respondent P9: *”CHPs hold monthly meetings at link facilities.”*

### Private sector integration challenges

Respondent P6: *”∼70% of immunizing facilities in County F are private/faith-based. Private facilities may be less likely to conduct outreach because of cost implications.”*

Respondent P5 (County E)”*Private facilities tend to have poorly trained staff - 2 FBOs [Faith based organisations] closed after KMPDC [Kenya Medical Practitioners and Dentists Council] inspection.”*

### Analysis of Thematic Findings through Complex adaptive System Theory

Table 3 maps the seven thematic findings from the deductive-inductive analysis onto six system-level constructs. These were also visualized in supplementary figure S2 according to their strength and direction of interactions.

**Table 3:** Translation of Themes to system-level constructs.

| Original Theme | System-level Construct | System-level Construct type |
| --- | --- | --- |
| Theme 1: Vaccine Procurement & Distribution | Supply Chain | Operational system element |
| Theme 2: Cold Chain Storage & Equipment | Supply Chain | Operational system element |
| Theme 3: Health Workforce | Health Workforce | Driver construct |
| Theme 4: Immunisation Financing | Financing | Exogenous constraint |
| Theme 5: Data and Records | Data Management | Feedback construct |
| Theme 6: Communication | Communication | Interaction/coupling construct |
| Theme 7: Service Delivery | Service Delivery | Resultant outcome |

### System-level Constructs

#### Supply Chain components: (Procurement + distribution + cold chain)

The supply chain represents the system’s capacity to make vaccines available at the point of use. It has 3 tightly coupled sub-components: Procurement -> Distribution -> Cold chain -> point of use.

The supply chain sub-system self-organises through redistribution between facilities, public transport use and drone delivery when formal pathways fail. The supply chain is highly sensitive to financing.

#### Financing as the exogenous constraint

Financing operates as an exogenous constraint that determines how effectively the other system-components function. It is highly influenced by political decisions and priority setting at county and national levels; as well as partner funding. The reliance of partner financing to meet domestic funding gaps creates a vicious cycle where the system adapts to partner presence rather than building domestic capacity.

The impact of the Social Health Authority (SHA) health financing model is beneficial in some counties and problematic in others.

Integration of immunization outreach activities with better funded programmes (HIV/TB) is a self-organising response to financing scarcity.

#### Health Workforce as a Driver of Service Delivery

The Health Workforce is the primary driver construct that transforms system inputs (vaccines, data tools and communication) into the system output (service delivery).

Single-nurse facilities represent a system at the “edge of chaos” where the smallest perturbation e.g. absence tips the system from functional to non-functional.

Health workers demonstrate remarkable self-organisation: using personal phones for communication, leveraging the Community Health Strategy and redistributing vaccines.

Training workshops pull the sole nurse away, paradoxically destroying the system instead of improving it. This is a CAS non-linearity where a positive intervention temporarily produces a short-term negative outcome.

#### Communication as a Cross-Cutting Interaction Construct

Communication mediates interactions between all the other system constructs. It is the coupling mechanism that determines how tightly or loosely the system’s components are connected. It enables and constrains the behaviour of every other construct.

The Kenya immunization system exhibits both formal (email cascade from the national to facility-level) and informal communication channels (phone messaging, personal phone calls and community radio). The informal pathways have emerged as adaptive responses to the rigidity and slowness of formal channels.

Vaccine hesitancy is the most important emergent property mediated by communication. We found out that hesitancy is predominantly antigen and class specific as exemplified by “elite” resistance to oral polio vaccine in one count, and present even among health care workers. This demonstrates that hesitancy is a result of complex interactions between communication and social context.

#### Data management as a Feedback Loop construct

Data management is the feedback loop construct that connects service delivery outcomes back to planning and resource allocation decisions. It represents the system’s capacity for learning and adaptation. Well-functioning feedback loops allow the system to self-correct; when they are broken, the system operates blind.

Redistribution of vaccines between facilities masks supply chain dysfunction in official data. The system’s data outputs do not reliably represent its actual state.

The monthly reporting cycle (15^th^ -25^th^) creates a temporal lag between events and system awareness. In a rapidly changing system, delayed feedback means interventions are always responding to past dates.

Data quality is directly determined by the same workforce that delivers services. When the sole nurse is overwhelmed, data quality is degraded. This creates a

coupling where the component most needed for system correction (data) degrades precisely when correction is needed (pressure on the staff and services).

#### Service delivery as the System Output Construct

Service delivery is the emergent output of the system and emerges from the interactions among all system-level constructs. This function is non-linear: doubling the vaccine supply does not double the coverage if the workforce cannot administer the vaccines. Improving data systems is of no benefit if there is inadequate workforce to act on the data. The system exhibits strong complementarities where all inputs must reach minimum thresholds simultaneously for the output to improve.

### Feedback Loops in the Kenya Immunisation System

Supplementary figure 3 illustrates the emergent reinforcing and balancing loops that influence immunisation service delivery.

#### Reinforcing loop (R1)

inadequate financing → poor supply → low coverage → low political priority → inadequate financing. Low financing leads to poor supply chain with consequent low coverage which erroneously presents immunization as a low priority health issue among the political stake-holders, further reducing budget allocation. This is a vicious cycle (positive feedback amplifying decline).

#### Reinforcing loop (R2)

Partner support → Reduced domestic investment → More partner dependency. External partner support may paradoxically reduce counties’ incentive to budget for immunization, creating a dependency trap.

#### Reinforcing loop (R3)

Poor working conditions→ staff attrition → fewer staff→ Increased workload → Poor working conditions. Poor working conditions lead staff to look for opportunities elsewhere, this worsens the existing staffing gap leading to increased workload. Reinforcing loop (R4): Vaccine redistribution → masked stockouts → false adequacy signal → no corrective action. Redistribution of vaccines between facilities masks stockouts and gives a false adequacy signal leading to no corrective action being taken.

#### Balancing loop (B1)

Coverage data → Targeted outreach → Improved coverage. KHIS data on low-coverage areas should trigger targeted outreach, correcting coverage gaps. However, this loop is weakened by data quality issues and workforce constraints.

#### Balancing loop (B2)

Stockout signals → E-Chanjo ordering → Resupply: E-Chanjo ordering is intended to match supply to demand. However, platform downtime and forecasting errors weaken this corrective mechanism.

#### Balancing loop (B3)

Vaccine Hesitancy → Low uptake observed → Community engagement →trust building. Vaccine hesitancy leads to low community trust. With community engagement, there is trust building.

#### Balancing loop (B4)

Service gaps → SHA budget allocation → improved funding →better service delivery. Service gaps are created by low funding. This is partly addressed by SHA budget allocation which improves funding leading to better service delivery.

#### Critical finding

While the system has equal numbers of reinforcing (R1-R4) and balancing loops (B1-B4), the reinforcing loops are more dominant and active than the balancing loops. This means the system tends towards amplifying dysfunction rather than self-correcting. The balancing loops that should provide stability (B1: coverage correction, B2: supply matching) are weakened by data quality problems and workforce constraints. This imbalance explains why the system persists in a sub-optimal state despite repeated interventions.

### Leverage points (Meadows’ Framework)

Using the Donella Meadows’ system theory we found that most common Kenya immunisation interventions focus on low-level operational changes such as budgets, staffing and stock capacity. While these may improve performance temporarily, complex adaptive systems theory suggests that the system’s reinforcing dynamics may eventually erode these gains. On the other hand, deeper interventions targeting paradigms, goals, governance rules, self-organisation and information flows have greater potential to create sustainable and long-term transformation because they change the underlying structure and behaviour of the system (Table 5).

**Table 5:**
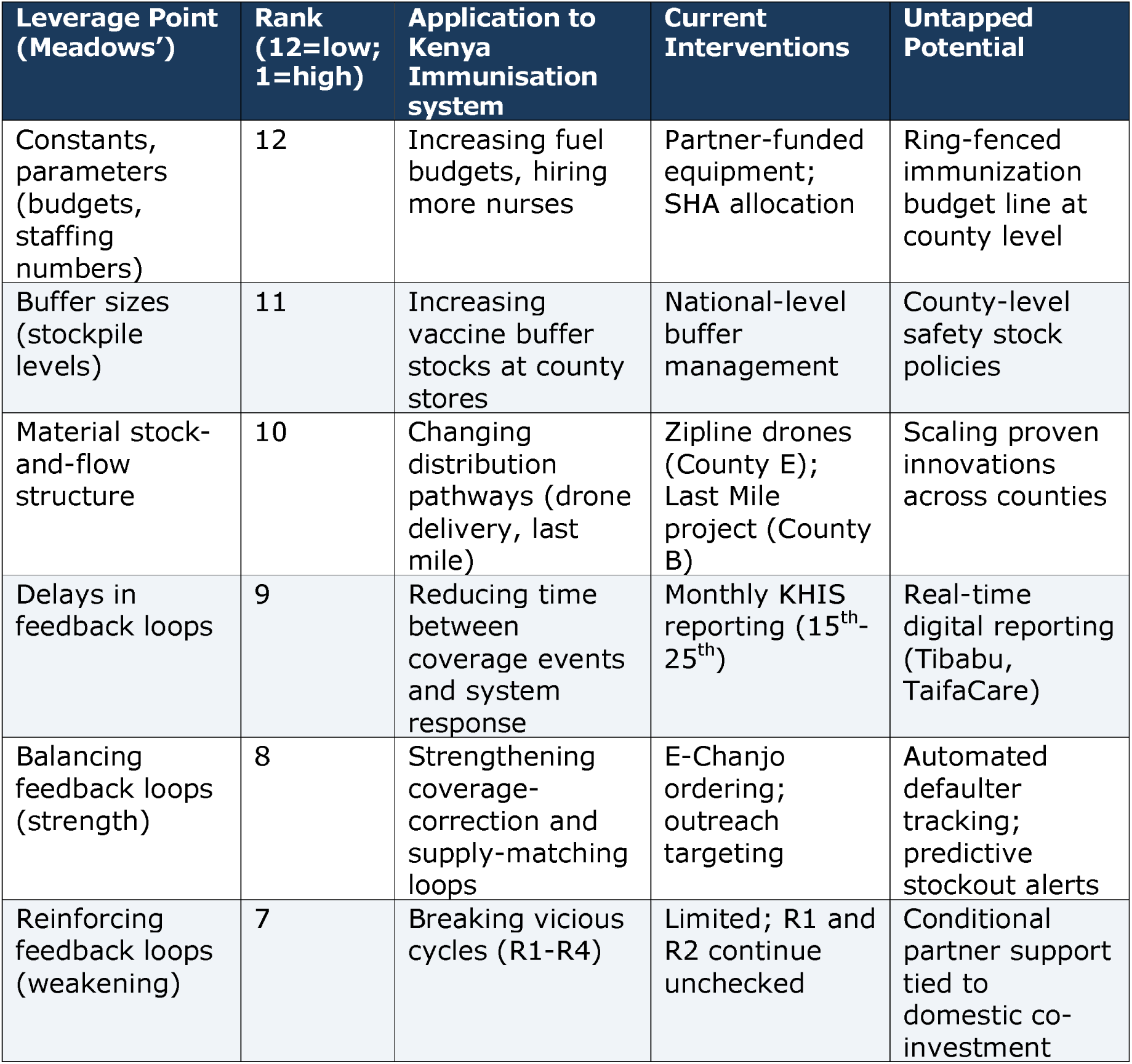
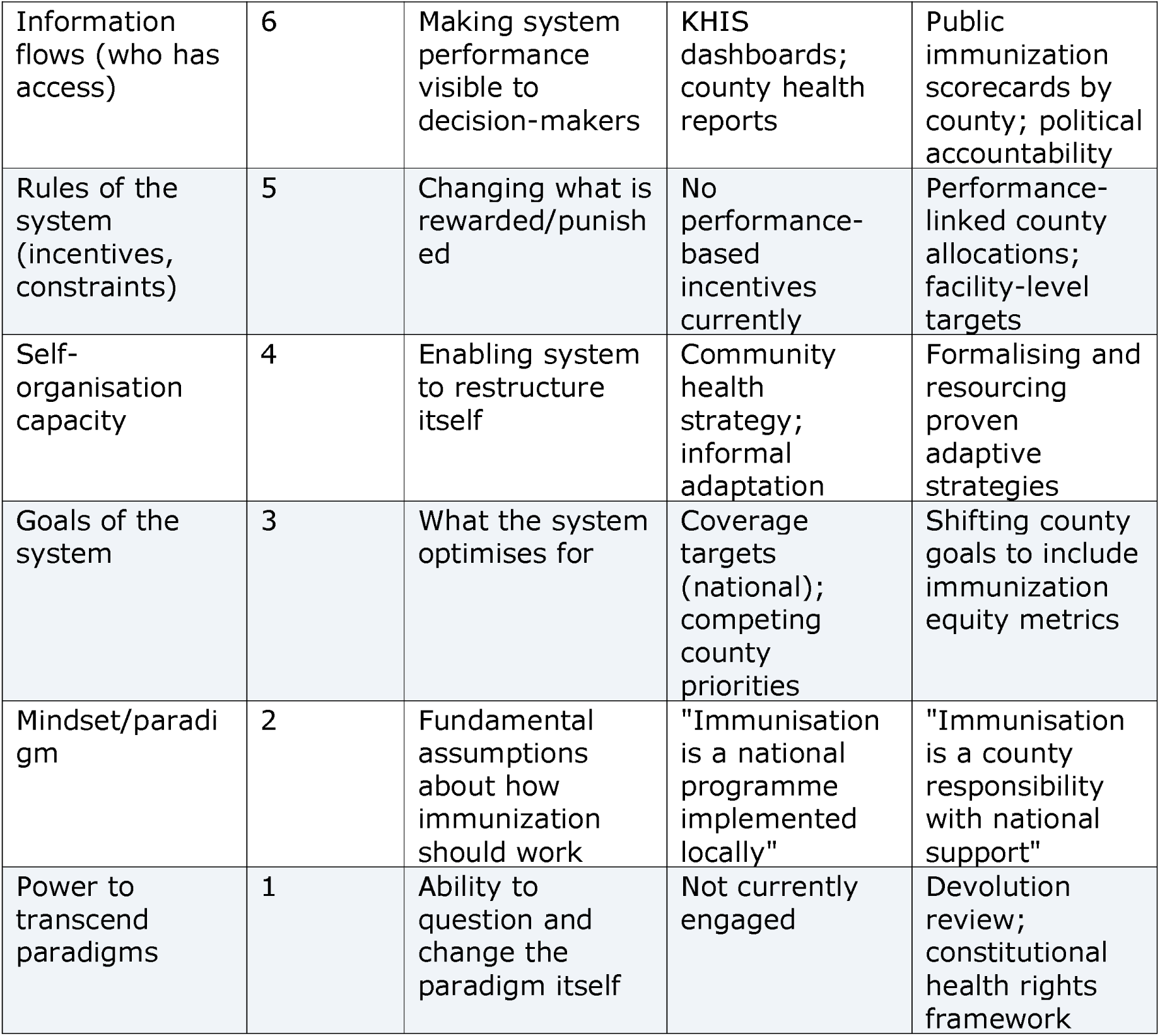
Meadows leverage points applied to the Kenya Immunisation system.

| Leverage Point (Meadows') | Rank (12=low; 1=high) | Application to Kenya Immunisation system | Current Interventions | Untapped Potential |
| --- | --- | --- | --- | --- |
| Constants, parameters (budgets, staffing numbers) | 12 | Increasing fuel budgets, hiring more nurses | Partner-funded equipment; SHA allocation | Ring-fenced immunization budget line at county level |
| Buffer sizes (stockpile levels) | 11 | Increasing vaccine buffer stocks at county stores | National-level buffer management | County-level safety stock policies |
| Material stock-and-flow structure | 10 | Changing distribution pathways (drone delivery, last mile) | Zipline drones (County E); Last Mile project (County B) | Scaling proven innovations across counties |
| Delays in feedback loops | 9 | Reducing time between coverage events and system response | Monthly KHIS reporting (15<sup>th</sup>-25<sup>th</sup>) | Real-time digital reporting (Tibabu, TaifaCare) |
| Balancing feedback loops (strength) | 8 | Strengthening coverage-correction and supply-matching loops | E-Chanjo ordering; outreach targeting | Automated defaulter tracking; predictive stockout alerts |
| Reinforcing feedback loops (weakening) | 7 | Breaking vicious cycles (R1-R4) | Limited; R1 and R2 continue unchecked | Conditional partner support tied to domestic co-investment |
| Information flows (who has access) | 6 | Making system performance visible to decision-makers | KHIS dashboards; county health reports | Public immunization scorecards by county; political accountability |
| Rules of the system (incentives, constraints) | 5 | Changing what is rewarded/punished | No performance-based incentives currently | Performance-linked county allocations; facility-level targets |
| Self-organisation capacity | 4 | Enabling system to restructure itself | Community health strategy; informal adaptation | Formalising and resourcing proven adaptive strategies |
| Goals of the system | 3 | What the system optimises for | Coverage targets (national); competing county priorities | Shifting county goals to include immunization equity metrics |
| Mindset/paradigm | 2 | Fundamental assumptions about how immunization should work | "Immunisation is a national programme implemented locally" | "Immunisation is a county responsibility with national support" |
| Power to transcend paradigms | 1 | Ability to question and change the paradigm itself | Not currently engaged | Devolution review; constitutional health rights framework |

### Implications for Policy and intervention design

The Meadows’ theory reframes how interventions should be designed for the Kenya immunisation system. Rather than linear, top-down solutions, the analysis identifies the following opportunities for the Kenyan context:

### Opportunity 1: intervene at higher leverage points

Current interventions cluster at shallow leverage points 10-12 which focus on changing quantities and resources (budgets, staffing, buffer stocks). Priority should shift to deeper level points 5-8 that involve changing system rules such as a shift to performance-based financing, strengthening information flows and weakening reinforcing vicious loops e.g. conditional partner support.

### Opportunity 2: Strengthen balancing loops before adding resources

Adding vaccines, equipment, or even staff will produce temporary improvements that erode if the balancing feedback loops (data → targeting → correction) are not strengthened first. Priority: fix the data feedback loop by investing in digital reporting, data quality, and workforce capacity to use data for local decisions.

### Opportunity 3: Formalise and resource self-organising behaviours

The system’s most effective adaptations (community health strategy, vaccine redistribution, phone messaging coordination, peer educators) emerged spontaneously. Rather than replacing these with formal top-down structures, policy should recognise, resource, and protect them.

### Opportunity 4: Design for heterogeneity, not uniformity

A systems-based policy would allow counties to adapt national strategies to local conditions while maintaining core performance standards. This flexibility is important because counties have unique environments and system constraints requiring context specific solutions.

### Opportunity 5: Break reinforcing vicious loops

The four identified vicious loops (R1: financing, R2: partner dependency, R3: workforce drain, R4: data masking) must be explicitly targeted. Each requires a different intervention: R1 needs ring-fenced budgets; R2 needs conditionality or rules to incentivise domestic financing; R3 needs competitive retention packages; R4 needs data audit and reconciliation protocols.

### Opportunity 6: Invest in system redundancy at the facility level

Single-nurse facilities operate at the edge of chaos with zero redundancy. The minimum viable intervention is ensuring every immunizing facility has at least two health workers. This is the single most impactful structural change to prevent the binary collapse dynamic.

### Opportunity 7: Treat Communication as infrastructure, not an add-on

Communication is the system’s coupling mechanism. Hesitancy, coordination failures, and training gaps are all fundamentally communication problems.

Investment in communication infrastructure (community radio, digital platforms, community health volunteer networks) should be treated as core system infrastructure.

## Discussion

Kenya’s immunisation performance is not simply a summation of individual determinants, but emerges from the interactions, feedback mechanisms and adaptive behaviours within several components of the system. Consequently, interventions targeting isolated components may have limited impact, and in some instances even generate unintended consequences [10, 17].

In this study, we explored perspectives of EPI personnel on immunisation program performance across selected counties and sub-counties in Kenya. Findings from thematic analysis were subsequently interpreted through complex adaptive systems and Meadows’ leverage points frameworks to examine how the determinants interact, how the system adapts to constraints, and where interventions have the potential to generate lasting change.

Out of the seven thematic areas explored, three themes emerged as prominent determinants of immunisation performance: vaccine distribution from the regional and county depots to immunising facilities, health workforce capacity and immunisation financing. Constraints affecting these system components were reported across all participating counties and sub-counties.

These findings are consistent with a case study analysis by Sakas et al in 2023, examining critical success factors in vaccine delivery across 3 countries (Nepal, Senegal and Zambia) with sustained high vaccination coverage. They showed that immunisation outcomes were dependent on political will, health-system strengthening, a robust community health strategy and adaptation of locally developed interventions [18]. Contextual adaptation has also been demonstrated by Decouttere and colleagues who, in 2023 applied a systems-thinking approach in studying immunisation sub-systems in Rwanda and Kenya. Through the participation of key EPI stakeholders, they developed a conceptual framework that identified root causes of poor performance and suggested possible directions for national immunisation system redesign for optimal service delivery [19].

In 2014, Varghese and colleagues [20] applied CAS theory to explore hidden dynamics contributing to declining vaccine coverage in Kerara, India. Their study identified several feedback loops involving changing actors and interactions that influenced vaccination decisions. Similar patterns of interconnectedness, adaptation and emergent behaviour were observed in the present study.

Regarding distribution of vaccines across counties, transport was identified as a major constraint, with respondents reporting lack of vehicles specifically designated for EPI activities, inadequate fuel budgets, long distances between vaccine stores and immunising facilities, difficult terrain and poor road networks. This illustrates how vaccine availability does not necessarily translate to successful vaccination if the vaccine does not reach the point of service delivery. To manage these transport constraints, counties adopted innovative solutions to maintain vaccine availability, including use of public transport, and the use of drones to transport vaccines to immunising facilities and outreach posts as reported in one county. This demonstrates that when formal structures fail, local actors usually develop solutions in an effort to sustain performance [18].

Health-worker shortages adversely affected immunisation performance. Many lower-level facilities reportedly depended on a single nurse responsible for clinical services, immunisation and data management. Consequently, excessive workload compromised service delivery and the quality of immunisation data, and when the sole nurse was absent, the facility was rendered non-operational forcing care-givers to seek services in more distant facilities. These findings augment evidence from a recent multi-country African study that identified staff shortages, high workloads and poor renumeration as important constraints to vaccine service delivery and data management. Competing service-delivery demands resulted in delayed data entry with a build up of data back logs [21].

Nevertheless, the workforce component demonstrated self-organisation and adaptation, with community-level structures supporting immunisation activities such as social mobilisation and defaulter tracing where facility-level human resource was inadequate. This task-sharing provides a temporary degree of functional redundancy in a fragile sub-system, and should be strengthened as the workforce deficit is being addressed. Leverage of the Community Health Strategy was also demonstrated in Malawi by Alban and colleagues in 2023. Health Surveillance Assistants (HSAs) in Malawi have an expanded scope of practice where they fully conduct routine immunisation activities. This task-shifting modality, though allowing continuity of essential immunisation services, may not be optimal as there is inadequate supervision support which raises safety concerns [22].

Insufficient immunisation financing was another prominent determinant of programme performance. Respondents reported declining partner funding and inadequate resources for EPI activities such as outreach and supervision.

Counties mitigated this challenge by combining immunisation activities with better funded programmes such as TB and HIV. These findings align with those of Adjagba and colleagues who identified limited budget allocations, delayed disbursement, political interference and resource allocation as important contributors to low financing in 15 Kenyan counties [23]. At the time of conducting this study, Kenya had commenced the process of transition from Gavi support to full domestic financing for vaccines. Ojal et al in 2019 projected a substantial increase in domestic vaccine expenditure, calling for the need to expand Kenya’s vaccine budget to sustain vaccine delivery following the cessation of Gavi funding [24]. Recent analyses on immunisation sustainability in sub-saharan Africa were conducted by Muyemba and colleagues [25]. They exposed vulnerability across all major system constructs, suggesting that strengthening immunisation performance should extend beyond operational improvements to include strategies that enhance system resilience and reduce donor dependency.

Application of the Meadows’ leverage-points system to the Kenya Immunisation system identified possible intervention points ranked according to the magnitude of potential impact. Interventions that focus on changing system structures and rules such as immunisation financing models and organisational restructuring at facility and county levels are likely to lead to sustainable positive change.

Lasting impact could also result from increasing visibility of immunisation within the political sphere, allowing it to be viewed as a priority item in the county health agenda as demonstrated by Sakas et al in 2023 [18]. This calls for advocacy activities to influence key stakeholders. Self-organising and adaptive behaviours within the system should be strengthened through allocation of more resources and adaptation of the emergent innovative coping strategies identified. For example, funding for the community health strategy that supports immunisation service delivery should be increased even as counties seek to address the perennial health worker shortage.

## Policy and Practice implications

This study highlights the need to go beyond mechanistic approaches to evaluating and improving immunisation performance in Kenya and similar contexts. The findings suggest that systems thinking approaches are necessary for identifying service delivery bottlenecks, understanding system vulnerabilities and designing interventions that target higher-level leverage points capable of generating sustainable system-wide improvements.

## Strengths and limitations

This study demonstrated several strengths, including the incorporation of diverse perspectives from frontline EPI personnel, the use of a multi-framework analytical approach and a systems-oriented interpretation of immunisation service delivery and data management.

However, several limitations should be acknowledged. The findings are context specific to Kenya and may not be directly generalisable to other settings. The cross-sectional qualitative design limited the ability to capture dynamic system evolution over time. This study interviewed only EPI coordinators, providing a single-level perspective on system dynamics. A broader scope of stakeholders including community health workers, caregivers, policy makers and private sector providers could offer substantially different accounts of system dynamics, identifying more feedback loops and potential intervention leverage points.

## Recommendations for future research

Future research should consider longitudinal systems studies, evaluations of leverage-based interventions and comparative analyses across districts or countries to better understand how immunisations systems evolve and respond to system-level interventions over time.

## Data Availability

All data produced in the present work are contained in the manuscript

## References

1. Ministry of Health. Kenya National Immunization Policy Guidelines Nairobi: Ministry of Health; 2023.

2. World Health Organisation. Immunization Agenda 2030: A Global Strategy to Leave No One Behind. WHO; 2020.

3 World Health Organisation/UNICEF. WHO/UNICEF estimates of national immunization coverage (WUENIC): Kenya. 2024.

4. Mekbib MJ, Thuo E, Gichuki J, Limo S, Doris Ogolla, et al. Immunization coverage estimates among children aged 12–23 months: Experience from a Mathare sub-county, Kenya. Journal of Interventional Epidemiology and Public Health. 2026;9(2):56.

5. Allan S, Adetifa IMO, Abbas K. Inequities in childhood immunisation coverage associated with socioeconomic, geographic, maternal, child, and place of birth characteristics in Kenya. BMC Infect Dis. 2021;21(1):553.

6. Karanja-Chege C, Agweyu A, Were F, Boele van Hensbroek M, Ogallo W. Addressing the challenges of estimating the target population in calculation of routine infant immunization coverage in Kenya. PLOS Global Public Health. 2025;5(7): e0004298.

7 World Health Organisation. Assessing and Improving the Accuracy of Target Population Estimates for Immunization Coverage. 2015.

8. Maina I, Wanjala P, Soti D, Kipruto H, Droti B, Boerma T. Using health-facility data to assess subnational coverage of maternal and child health indicators, Kenya. Bulletin of the World Health Organization. 2017; 95:683–94.

9 Karanja-Chege CM, Jalang’o R, Agweyu A, Were F, Boele van Hensbroek M, Ogallo W. Assessing immunization system performance through coverage, dropout, and health facility density in Kenya from 2020-2024. Journal of Global Health Economics and Policy. 2026;6.

10. Carey G, Malbon E, Carey N, Joyce A, Crammond B, Carey A. Systems science and systems thinking for public health: a systematic review of the field. BMJ Open. 2015;5(12):e009002.

11. Meadows DH. Leverage points: Places to intervene in a system Hartland, VT: Sustainability Institute. 1999

12 World Health Organisation Africa Regional Office. Regional strategic plan for immunization 2014–2020. 2014.

13 World Health Organisation. Monitoring the building blocks of health systems: A handbook of indicators and their measurement strategies. 2010.

14. Braun V and Clarke V. Thematic Analysis: A Practical Guide. Sage 2021.

15 Microsoft Copilot [Large language model] [Internet]. 2026. Available from: https://copilot.microsoft.com/.

16. Tong A, Sainsbury P, Craig J. Consolidated criteria for reporting qualitative research (COREQ): a 32-item checklist for interviews and focus groups. Int J Qual Health Care. 2007;19(6):349–57.

17. Adam T, de Savigny D. Systems thinking for strengthening health systems in LMICs: need for a paradigm shift. Health Policy Plan. 2012;27 Suppl 4: iv1-3.

18. Sakas Z, Hester KA, Ellis A, Ogutu EA, Rodriguez K, Bednarczyk R, et al. Critical success factors for high routine immunisation performance: a qualitative analysis of interviews and focus groups from Nepal, Senegal, and Zambia. BMJ Open. 2023;13(10): e070541.

19. Decouttere C, Vandaele N, De Boeck K, Banzimana S. A Systems-Based Framework for Immunisation System Design: Six Loops, Three Flows, Two Paradigms. Health Systems. 2023;12(1):36–51.

20. Varghese J, Kutty VR, Paina L, Adam T. Advancing the application of systems thinking in health: understanding the growing complexity governing immunization services in Kerala, India. Health Research Policy and Systems. 2014;12(1):47.

21. Carnahan E, Van Grack A, Kangethe B, Diallo MM, Mutai D, Bah O, et al. Root causes of COVID-19 data backlogs: a mixed methods analysis in four African countries. Oxford Open Digital Health. 2024;2(Supplement_1): i16–i28.

22. Alban R, Gibson E, Payne J, Chihana T. Leveraging community health workers as vaccinators: a case study exploring the role of Malawi’s Health Surveillance Assistants in delivering routine immunization services. Human Resources for Health. 2023;21(1):42.

23. Adjagba AO, Oguta JO, Akoth C, Wambiya EOA, Nonvignon J, Jackson D. Financing immunisation in Kenya: examining bottlenecks in health sector planning and budgeting at the decentralised level. Cost Effectiveness and Resource Allocation. 2024;22(1):76.

24. Ojal J, Griffiths U, Hammitt LL, Adetifa I, Akech D, Tabu C, et al. Sustaining pneumococcal vaccination after transitioning from Gavi support: a modelling and cost-effectiveness study in Kenya. Lancet Glob Health. 2019;7(5): e644–e54

25. Myemba DT, Smets L, Sunguya BF, Vandaele N, Decouttere C. Challenges and strategies for sustainable and resilient immunization systems in sub-Saharan Africa: A comprehensive scoping review. Vaccine. 2025; 45:126639.

