## Supplementary material for "Perspectives from EPI personnel on immunisation programme performance in Kenya: a thematic, complex adaptive systems and leverage-points analysis": supplemmantary file

**Supplementary material 1:**

**Consent form**

**Study Title**: Perspectives from County EPI Key Personnel on Immunisation Service Delivery and Data Use

**PhD Candidate:** Dr. Christine Karanja-Chege, (MBChB, MMed Paediatrics and Childhealth, PgDip.Paediatric Infectious Diseases)

**Supervisors:**

1. Prof. Michaël Boele van Hensbroek (MD, PhD, Professor in Global Child Health)
2. Prof. Fred Were (MBChB, MMed Paediatrics and Childhealth, PhD, Professor of perinatal and neonatal medicine)
3. Dr. William Ogallo (B Pharm, PhD)

**Doctoral School:** Amsterdam University Medical Centers Doctoral School

You are invited to take part in a research study as a Key Informant interviewee. This form contains information that will help you decide whether to participate in the study.

**Key Information**

- This interview aims to explore the contextual factors that may account for the immunization performance patterns observed from the previous quantitative analyses across the counties.
- If you choose to participate, you will be asked to answer questions regarding immunization service delivery and data use.
- We do not anticipate any unusual risk to you as the information we expect from you is not directly regarding any individual and therefore there is minimal risk of a breach in confidentiality.
- The direct benefits of your participation are increased knowledge on the factors affecting immunization service delivery and data use in Kenya which may be of importance in guiding policy-makers in Kenya.
- Taking part in this research project is voluntary. You do not have to participate and you can stop at any time. Please take time to read this entire form and ask questions before deciding whether to take part in this research project.

**Financial Information**

You will not receive financial compensation for taking part in the study and you will also not be required to pay to be part of the study.

**Protecting and sharing research information**

The interview audio and transcript files will only be accessible to study team. Access to the database will be through a secure password.

We will not keep your name or other information that can identify you directly.

The results of this study will be published in an article or presentation, but will not include any information that would let others know who you are.

**Contact Information**

In case of any concerns or questions, please contact the researcher whose contacts are provided below:

**Principal Investigator: Christine Karanja-Chege**

****

*I understand what the study is about and my questions so far have been answered. I agree to take part in this study.*

Participant Name _____________________________________________

Signature__________________	Date __________________

**Supplementary Material 2:**

**Thematic Key Personnel Interview Guide**

| **Thematic Area** | **Prompt** | **Probes (with examples where possible)** |
| --- | --- | --- |
| **Vaccines & Commodities Procurement** | Describe how vaccines and commodities are procured in your county. | - How are quantities determined? Who are involved? Challenges with delays, shortages, quality? |
| **Distribution Process** | Explain how vaccines are distributed to health facilities. | - How is delivery tracked? Challenges in hard to reach areas?- Recent bottlenecks or successes? |
| **Storage & Cold Chain** | Tell me about storage facilities and cold chain equipment. | - Types of equipment and maintenance?- Issues maintaining temperature?- Equipment upgrades or repairs? |
| **Data & Records Management** | How is immunization data collected and managed? | - Accuracy and timeliness of reporting?- How is data used for planning?- Challenges with data entry or reporting? |
| **Health Workforce** | Describe the immunization workforce at the county level. | - Are staff numbers adequate?- Training and support provided?- Monitoring performance and accountability? |
| **Communication** | How do you communicate program info to workers and communities? | - Strategies for vaccine hesitancy?- Challenges in timely info dissemination? |
| **Immunization Financing** | Explain how immunization is financed in the county. | - Funding gaps or delays?- Resource allocation and sustainability? |
| **Service Delivery at County Level** | Describe how immunization services are organized. | - Outreach/mobile services?- Challenges in equitable access?- Notable successes or innovations? |
| **Closing** | Please add any other information that you would like about immunization in your county | - Key recommendations for improving immunization in your county? |

**Supplementary Figure S1:**

Coding tree of themes and sub-themes influencing immunization programme performance

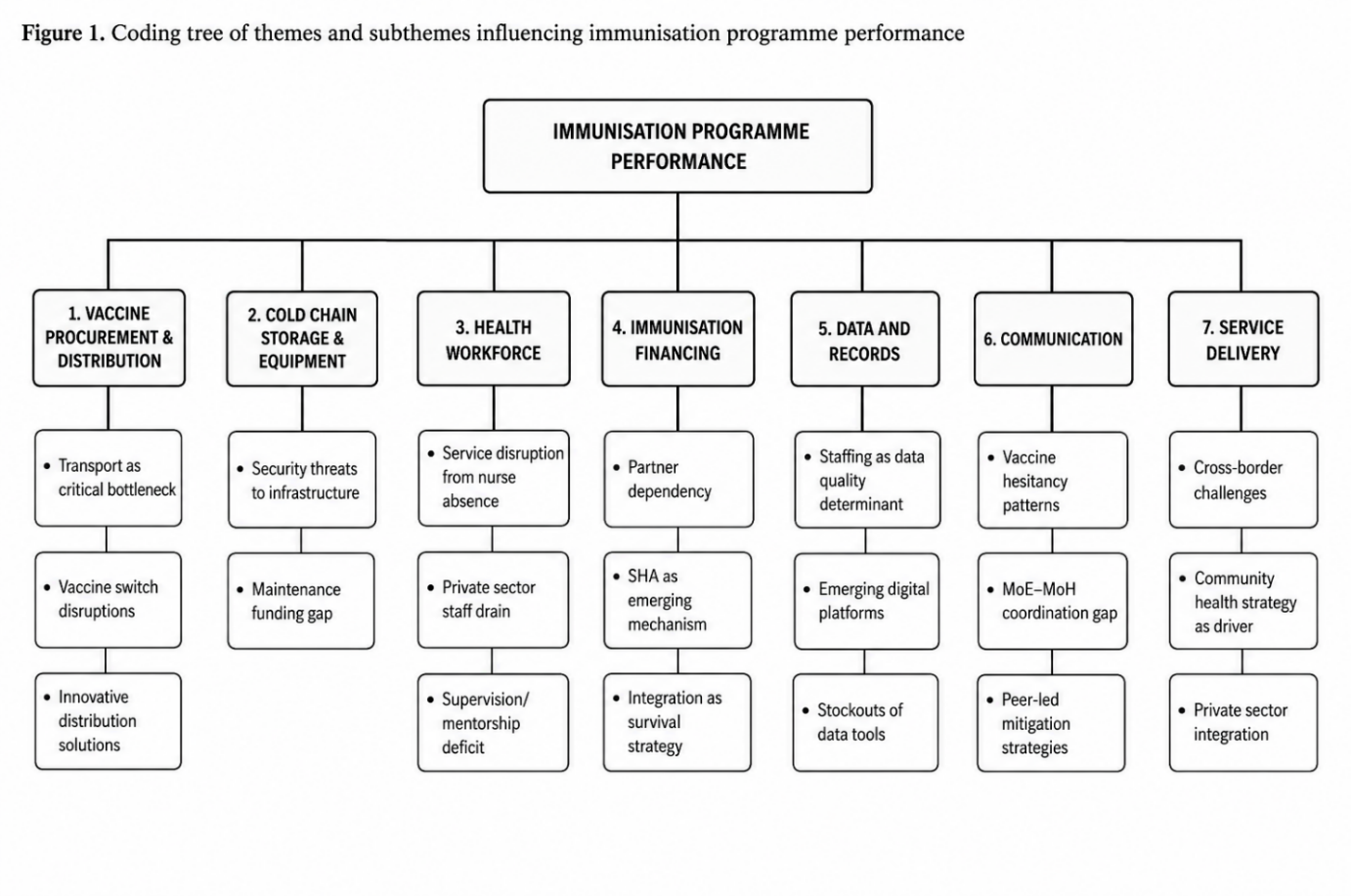

**Suppementary Figure 1:** Coding tree of themes and sub-themes influencing immunization programme performance

**Supplementary Figure S2:**

Directional interaction map of the six CAS constructs.

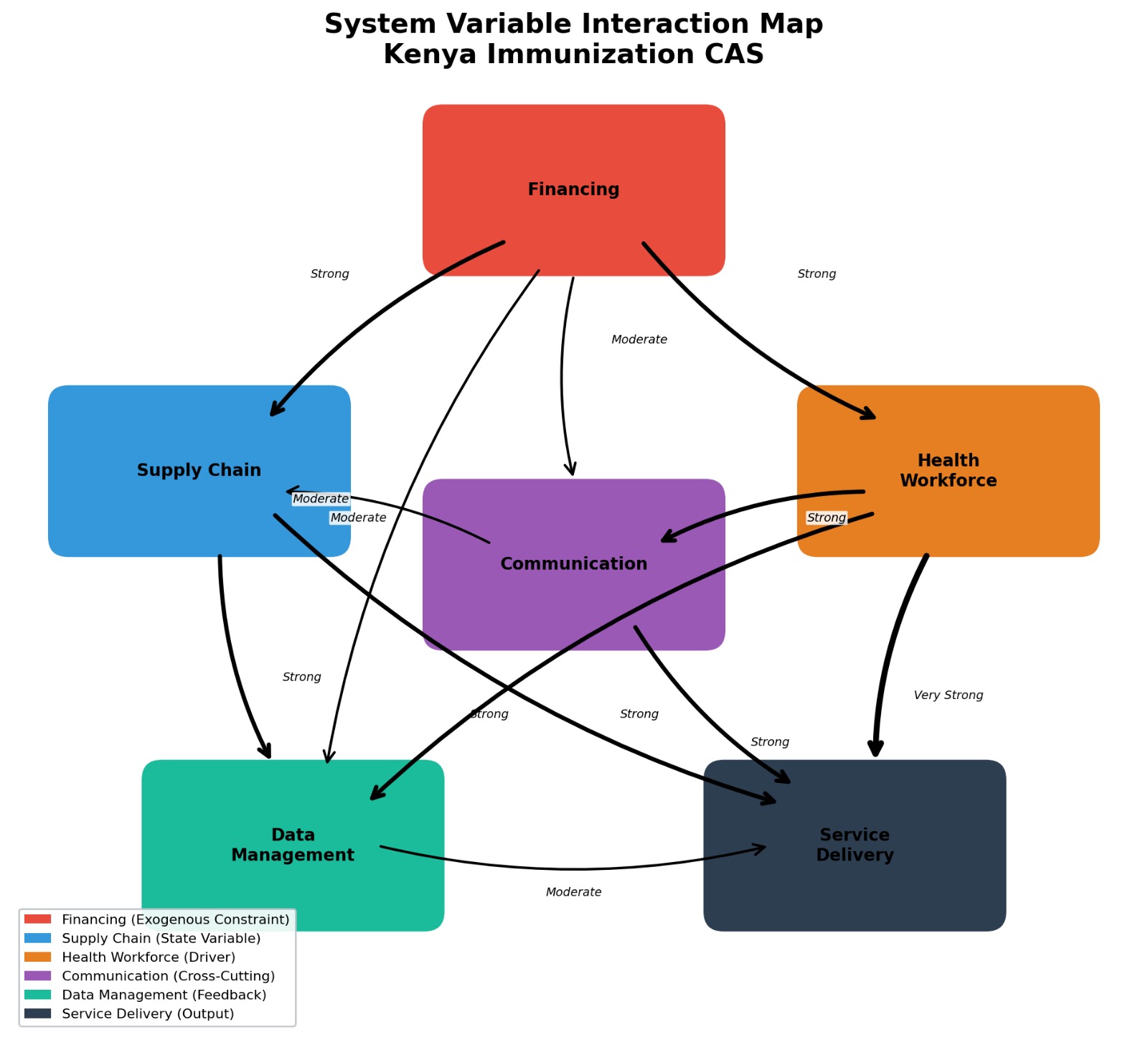

**Supplementary figure S2:** Directional interaction map of the 6 CAS variables. Arrow thickness and labels indicate the interaction strength. Financing is the most influential exogenous variable; Health Workforce has a “very strong” direct effect on Service Delivery

**Supplementary Figure S3:**

Feedback loops in Kenya’s immunization system.

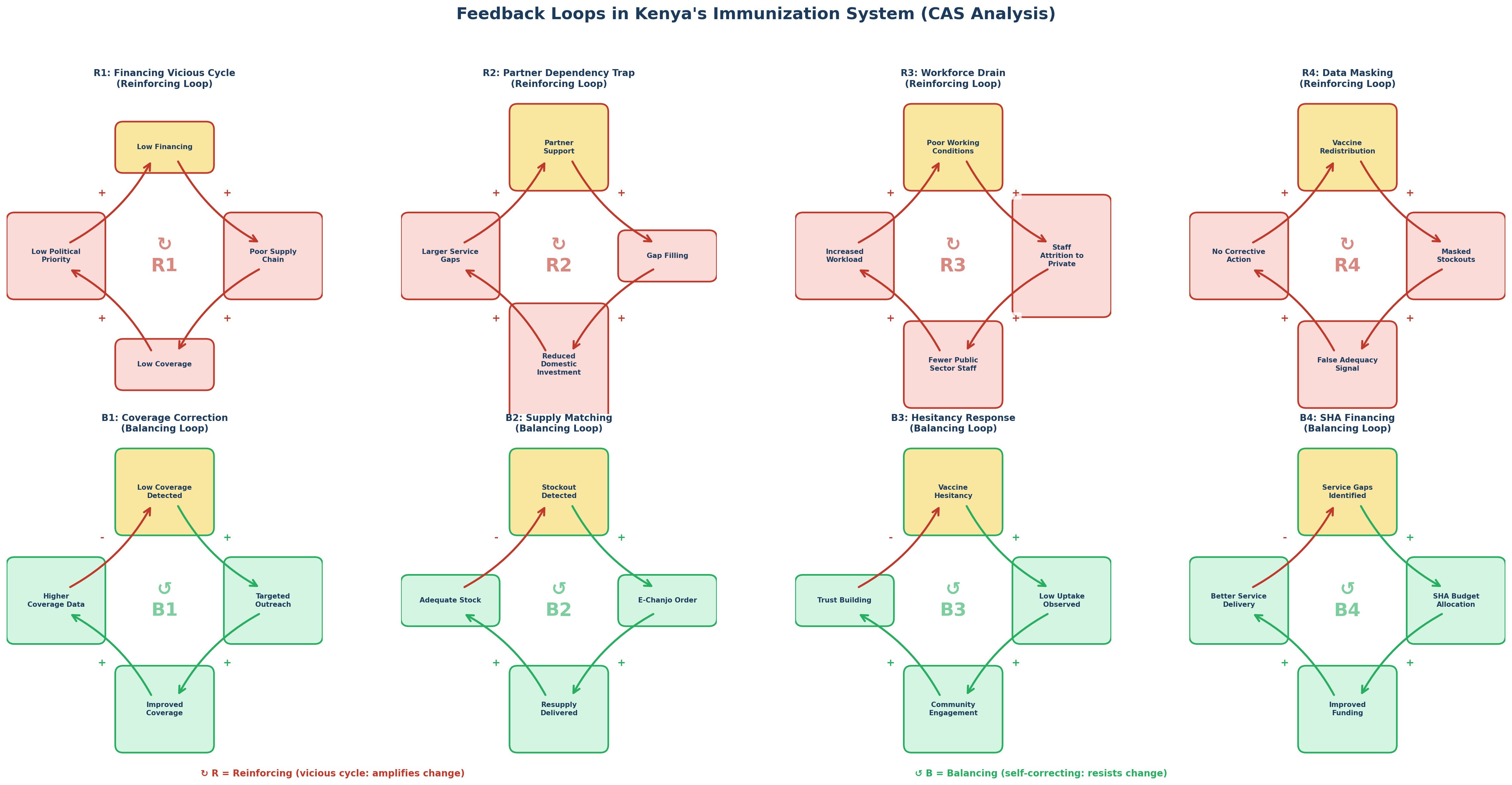

**Figure 3:** Feedback loops in Kenya’s immunization system.

The reinforcing loops (R1-R4) are more dominant and active than the effective balancing loops (B1-B4) tending to amplify system dysfunction
